# Impact of Pre-reperfusion Left Ventricle Unloading on ST-segment-elevation Myocardial Infarction According to the Onset-to-Unloading Time

**DOI:** 10.1101/2024.01.29.24301969

**Authors:** Naotaka Okamoto, Yasuyuki Egami, Masaru Abe, Mizuki Osuga, Hiroaki Nohara, Shodai Kawanami, Akito Kawamura, Kohei Ukita, Koji Yasumoto, Masaki Tsuda, Yasuharu Matsunaga-Lee, Masamichi Yano, Masami Nishino, J-PVAD investigators

## Abstract

**Background:** Impella in the left ventricle (LV) with delaying reperfusion reduces the infarction size in animal models. However, the onset-to-unloading time in real-world practice can frequently be longer than that in animal experiments. It is unclear whether the impact of pre-reperfusion Impella use is sustained throughout all periods from the onset. This study aimed to evaluate the association between the onset-to-unloading time and the effect of pre-reperfusion Impella on the survival of patients with ST-segment-elevation myocardial infarction (STEMI).

**Methods:** This study is a post-hoc analysis of the J-PVAD registry. Among all patients registered in J-PVAD between February 2020 and December 2021, patients with STEMI and treated with Impella alone support were selected. Two cohorts were provided based on whether the onset-to-unloading time was less than 6 hours. The patients were divided into two groups according to pre- or post-reperfusion use of Impella in each cohort. The primary outcome was an 80-day survival rate. The independent factors of survival were identified with a multivariable Cox proportional hazard regression analysis after adjusting for the variables that were statistically significant in the univariable analysis.

**Results:** Patients with pre-reperfusion unloading had a significantly higher 80-day survival rate than patients with post-reperfusion unloading (81.6% vs. 59.5%, p=0.02) in the cohort with an onset-to-unloading time ≥6 hours, while patients with pre- and post-reperfusion unloading had similar 80-day survival rates (85.3% vs. 91.2%, p=0.38) in the cohort with an onset-to-unloading time <6 hours. A multivariable analysis revealed that pre-reperfusion use of Impella was an independent factor of survival (hazard ratio 0.184 [95% confidence interval 0.045-0.746], p=0.02) in the onset-to-unloading time ≥6 hours cohort.

**Conclusions:** Pre-reperfusion LV unloading could be a crucial treatment to improve the short-term survival rate when the onset-to-LV unloading was more than 6 hours.

## Introduction

Primary percutaneous coronary intervention (PCI) is no doubt the gold-standard strategy for patients with ST-segment-elevation myocardial infarction (STEMI). Previous guidelines had emphasized achieving a door-to-balloon time of 90 minutes or less for many years ^1^, but several studies have shown no association between the door-to-balloon time and mortality in patients with STEMI ^2, 3^. It has been demonstrated that there is a stronger correlation between mortality and the onset-to-balloon time compared to the door-to-balloon time ^3^. Therefore, not only reperfusion with no delay but shortening the transfer time from the onset are highlighted by the current guidelines ^4, 5^. Contrary to the early reperfusion strategy, it has been reported that left ventricle (LV) unloading using a catheter-based micro-axial ventricular assist device Impella (Abiomed, Danvers, MA, USA) with delaying reperfusion reduces the infarction size in animal models ^6, 7^. There are many differences including the age, comorbidities, and presence of prodomal angina between animal models and real-world patients. In terms of the time from onset, a primary PCI is recommended within 12 hours of onset and is indicated in the presence of ongoing symptoms suggestive of ischemia, hemodynamic instability, or life-threatening arrhythmias even after 12 hours. That is to say, the onset-to-unloading time in real-world practice can frequently be longer than that in animal experiments, in which the onset-to-unloading time is mostly less than two hours. Registries of patients with myocardial infarction treated with Impella in the USA have revealed that Impella use prior to PCI is associated with an improved survival ^8, 9^. However, it is unclear whether the impact of the pre-reperfusion Impella use is sustained throughout all periods from the onset. This study aimed to evaluate the association between the onset-to-unloading time and the effect of pre-reperfusion with Impella on the survival of patients with STEMI.

## Methods

### Study population

This study is a post-hoc analysis of the J-PVAD registry. J-PVAD is an ongoing multicenter observational registry certified by the academic-based Japan Impella Committee and registered with the Universal Hospital Medical Information Network (UMIN) Clinical Trial Registry (ID: UMIN 000033603). All patients with acute heart failure and an attempted or successful placement of Impella 2.5, CP or 5.0 pump at qualified centers were enrolled in the J-PVAD. The study complied with the Declaration of Helsinki and was approved by the Central Institution Review Board at Osaka University. Individual patient data were stored directly in a centralized electronic database by participating investigators. Among all patients registered in the J-PVAD between February 2020 and December 2021, the patients with STEMI were selected. Patients treated with extracorporeal membrane oxygenation were excluded since the pure effect of pre-reperfusion Impella use could not be evaluated. Furthermore, patients without data regarding the onset-to-unloading time, timing of the Impella support (pre- or post-reperfusion), or survival data and patients who did not undergo PCI were excluded. Patients with STEMI who underwent PCI more than 6 hours after the onset have been reported to have a higher mortality rate than those who underwent PCI within 6 hours ^3^. Two cohorts were provided based on whether the onset-to-unloading time was less than 6 hours. The patients were divided into two groups according to pre- or post-reperfusion use of Impella in each cohort (Figure 1).

**Figure 1.**
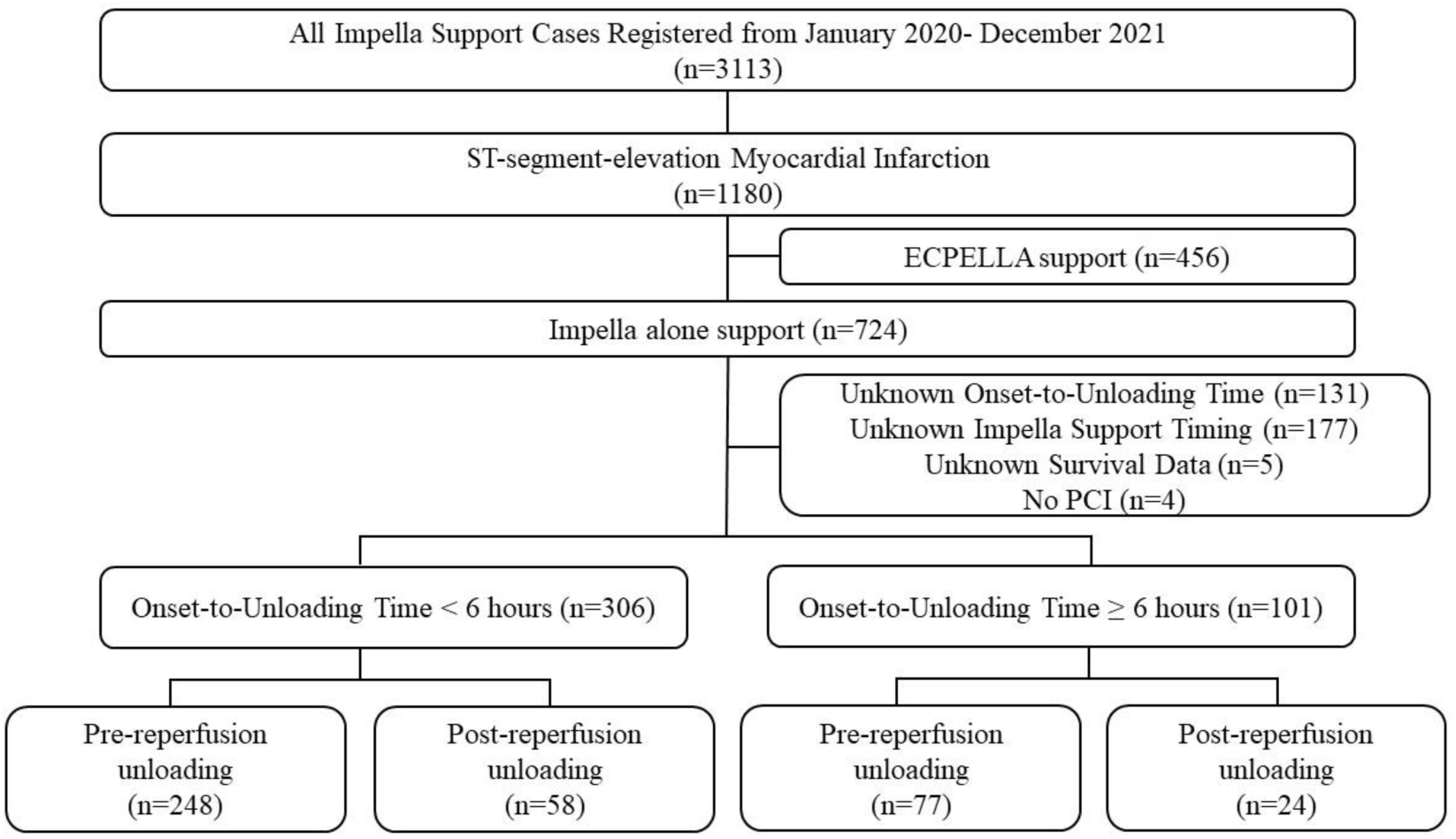
Study flow chart ECPELLA= Impella plus venoarterial extracorporeal membrane oxygenation

### Clinical outcomes

The survival rate at 80 days was evaluated in each group in the two cohorts. Furthermore, the factors associated with survival were evaluated in each cohort. Major adverse events including hemolysis, hemorrhage, hematoma, peripheral ischemia, and strokes were assessed. Hemolysis was defined as an event with an increase in the plasma free hemoglobin of more than 40 mg/dl or a clinically significant increase in the lactate dehydrogenase and indirect bilirubin and a decrease in the hemoglobin level within 48 hours after the Impella support. Hemorrhage was defined as bleeding which required surgical intervention or transfusion. Hematoma was defined as hematoma that was larger than 5 cm in diameter or required surgical intervention. Peripheral ischemia was defined as limb ischemia that was treated with medical or surgical intervention. Strokes were defined as a clinically significant neurological deficit or apparent infarction or/and intracranial hemorrhage ^10, 11^.

### Statistical analysis

Data are presented as the mean ± standard deviation or the median (25th percentile-75th percentile) and were compared by a *t-*test or the Kruskal-Wallis test according to the distribution. Categorical variables are presented as numbers and percentages and were compared by either the chi-square test or Fisher’s exact test as appropriate. A Kaplan-Meier estimate with a log-rank test was used to compare the survival rates among patients who underwent pre- or post-reperfusion Impella support. Predictors of survival were assessed from the patient characteristics, laboratory data, and procedure characteristics in a univariable Cox proportional hazard regression analysis. The independent factors of survival were identified with a multivariable Cox proportional hazard regression analysis after adjusting for the variables that were statistically significant in the univariable analysis. A value of P<0.05 was considered to be statistically significant. Statistical analyses were performed using JMP version 17.0.0 software (SAS Institute Inc., Cary, North Carolina, USA).

## Results

### Patient characteristics

The patient characteristics are summarized in Table 1. In the cohort with an onset-to-unloading time <6 hours, the age, gender, and prevalence of comorbidities did not differ between the groups. The heart rate, systolic and diastolic blood pressure before the Impella insertion, and ejection fraction were also similar. The creatine kinase and C-reactive protein were significantly greater in the patients with pre-reperfusion unloading than in those with post-reperfusion unloading. The other laboratory data did not differ between the two groups. In the cohort with an onset-to-unloading time ≥6 hours, there was no difference between the two groups.

**Table 1.** Patient characteristics.

|  | Door-to-Unloading Time <6 hours |  |  | Door-to-Unloading Time ≥6 hours |  |  |
| --- | --- | --- | --- | --- | --- | --- |
|  | Pre-reperfusion group<br>(n=248) | Post-reperfusion<br>group (n=58) | P-value | Pre-reperfusion group<br>(n=77) | Post-reperfusion group<br>(n=24) | P-value |
| Age, number assessed, n (%) | 248 (100) | 58 (100) |  | 76 (98.7) | 24 (100) |  |
| Age, year | 70.0±11.8 | 67.2±11.8 | 0.11 | 71.8±11.3 | 72.3±12.2 | 0.86 |
| Gender, number assessed, n (%) | 248 (100) | 58 (100) |  | 77 (100) | 24 (100) |  |
| Male, n (%) | 209 (84) | 50 (86) | 0.85 | 62 (81) | 17 (71) | 0.39 |
| Body mass index, number assessed, n (%) | 242 (97.6) | 56 (96.6) |  | 75 (97.4) | 23 (95.8) |  |
| Body mass index, kg/m<sup>2</sup> | 23.4±3.7 | 23.2±3.8 | 0.80 | 22.4 ±3.6 | 23.4±3.9 | 0.29 |
| Hypertension, number assessed, n (%) | 243 (98.0) | 58 (100) |  | 77 (100) | 24 (100) |  |
| Hypertension | 167 (69) | 34 (59) | 0.16 | 49 (64) | 20 (83) | 0.08 |
| Dyslipidemia, number assessed, n (%) | 245 (98.8) | 58 (100) |  | 77 (100) | 24 (100) |  |
| Dyslipidemia | 133 (54) | 34 (59) | 0.48 | 37 (48) | 11 (46) | 1.00 |
| Diabetes mellitus, number assessed, n (%) | 244 (98.4) | 58 (100) |  | 76 (98.7) | 24 (100) |  |
| Diabetes mellitus | 103 (42) | 20 (34) | 0.30 | 32 (42) | 14 (58) | 0.24 |
| Chronic kidney disease, number assessed, n (%) | 241 (97.2) | 56 (96.6) |  | 77 (100) | 23 (95.8) |  |
| Chronic kidney disease | 69 (29) | 15 (27) | 0.87 | 19 (25) | 8 (35) | 0.42 |
| Out-hospital cardiac arrest, number assessed, n (%) | 247 (99.6) | 58 (100) |  | 77 (100) | 24 (100) |  |
| Out-hospital cardiac arrest | 26 (11) | 3 (5) | 0.32 | 6 (6) | 1 (4) | 1.00 |
| Heart rate before Impella insertion, number assessed, n (%) | 247 (99.6) | 57 (98.3) |  | 77 (100) | 22 (91.7) |  |
| Heart rate before Impella insertion, bpm | 89.2±26.2 | 90.1±26.0 | 0.81 | 96.8±22.0 | 95.1±20.5 | 0.73 |
| Systolic blood pressure before Impella insertion, number assessed, n (%) | 246 (99.2) | 57 (98.3) |  | 77 (100) | 23 (95.8) |  |
| Systolic blood pressure before Impella insertion, mmHg | 103.9±20.2 | 98.8±17.3 | 0.26 | 102.7±25.2 | 99.1±26.3 | 0.57 |
| Diastolic blood pressure before Impella insertion, number assessed, n (%) | 247 (99.6) | 57 (98.3) |  | 77 (100) | 22 (91.7) |  |
| Diastolic blood pressure before Impella insertion, mmHg | 67.7±23.6 | 62.4±19.1 | 0.07 | 66.5±20.7 | 62.8±20.3 | 0.46 |
| Ejection fraction, number assessed, n (%) | 119 (48.0) | 24 (41.4) |  | 45 (58.4) | 19 (79.2) |  |
| Ejection fraction, % | 30 (20-40) | 30 (27-45) | 0.42 | 30 (23-40) | 30 (23-38) | 0.99 |
| Lactate, number assessed, n (%) | 180 (72.6) | 33 (56.9) |  | 59 (76.6) | 14 (58.3) |  |
| Lactate, mmol/l | 4.10 (2.31-6.11) | 3.30 (2.55-5.05) | 0.28 | 2.5 (1.4-5.0) | 3.4 (1.6-4.9) | 0.97 |
| Lactate dehydrogenase, number assessed, n (%) | 233 (94.0) | 55 (94.8) |  | 72 (93.5) | 24 (100) |  |
| Lactate dehydrogenase, U/L | 258 (200-368) | 234 (189-280) | 0.04 | 404 (289-667) | 380 (249-791) | 0.68 |
| Creatinine, number assessed, n (%) | 248 (100) | 58 (100) |  | 76 (98.7) | 24 (100) |  |
| Creatinine, mg/dl | 1.09 (0.87-1.35) | 0.96 (0.88-1.21) | 0.45 | 1.01 (0.81-1.39) | 1.26 (0.86-1.52) | 0.16 |
| Total bilirubin, number assessed, n (%) | 235 (94.8) | 54 (93.1) |  | 71 (92.2) | 24 (100) |  |
| Total bilirubin, mg/dl | 0.6 (0.5-0.8) | 0.6(0.5-0.9) | 0.23 | 0.8 (0.6-1.0) | 0.8 (0.5-1.0) | 0.34 |
| Albumin, number assessed, n (%) | 234 (94.4) | 53 (91.4) |  | 71 (92.2) | 23 (95.8) |  |
| Albumin, g/dl | 3.9 (3.6-4.2) | 4.0 (3.7-4.2) | 0.09 | 3.6 (3.2-4.0) | 3.3 (2.8-4.0) | 0.09 |
| Aspartate aminotransferase, number assessed, n (%) | 242 (97.6) | 56 (96.6) |  | 74 (96.1) | 24 (100) |  |
| Aspartate aminotransferase, U/L | 36 (24-66) | 30 (21-46) | 0.07 | 108 (52-235) | 72 (34-168) | 0.21 |
| Alanine aminotransferase, number assessed, n (%) | 242 (97.6) | 56 (96.6) |  | 74 (96.1) | 24 (100) |  |
| Alanine aminotransferase, U/L | 26 (17-45) | 23 (14-46) | 0.28 | 36 (21-74) | 35 (14-46) | 0.17 |
| Creatine phosphokinase, number assessed, n (%) | 247 (99.6) | 58 (100) |  | 77 (100) | 24 (100) |  |
| Creatine phosphokinase, U/L | 140 (90-281) | 111 (74-177) | 0.02 | 665 (346-1889) | 529 (225-1264) | 0.31 |
| C-reactive protein, number assessed, n (%) | 233 (90.3) | 56 (96.6) |  | 73 (94.8) | 24 (100) |  |
| C-reactive protein, mg/dl | 0.14 (0.06-0.54) | 0.10 (0.03-0.25) | 0.04 | 0.67 (0.11-3.67) | 2.29 (0.33-6.49) | 0.09 |

1 Values are the mean ± standard deviation, median (interquartile) or n (%).

### Procedural characteristics

Table 2 shows the procedural characteristics. In the cohort with an onset-to-unloading time <6 hours, the onset-to-unloading and door-to-unloading time were significantly shorter in the pre-reperfusion unloading group than in the post-reperfusion unloading group (150 [111-210] vs. 192 [156-260] minutes, p<0.01, 72 [56-100] vs. 104 [80-134] minutes, p<0.01, respectively). The door-to-balloon time was conversely longer in the pre-reperfusion unloading group than in the post-reperfusion unloading group (91 [71-127] vs. 59 [45-92] minutes, p<0.01). On the other hand, in the cohort with an onset-to-unloading time≥ 6 hours, the onset-to-unloading, door-to-unloading, and door-to-balloon time did not differ between the pre- and post-reperfusion unloading groups. Inotropes were used in most of the patients and more than 90% of the patients were treated with Impella CP in all groups.

**Table 2.**
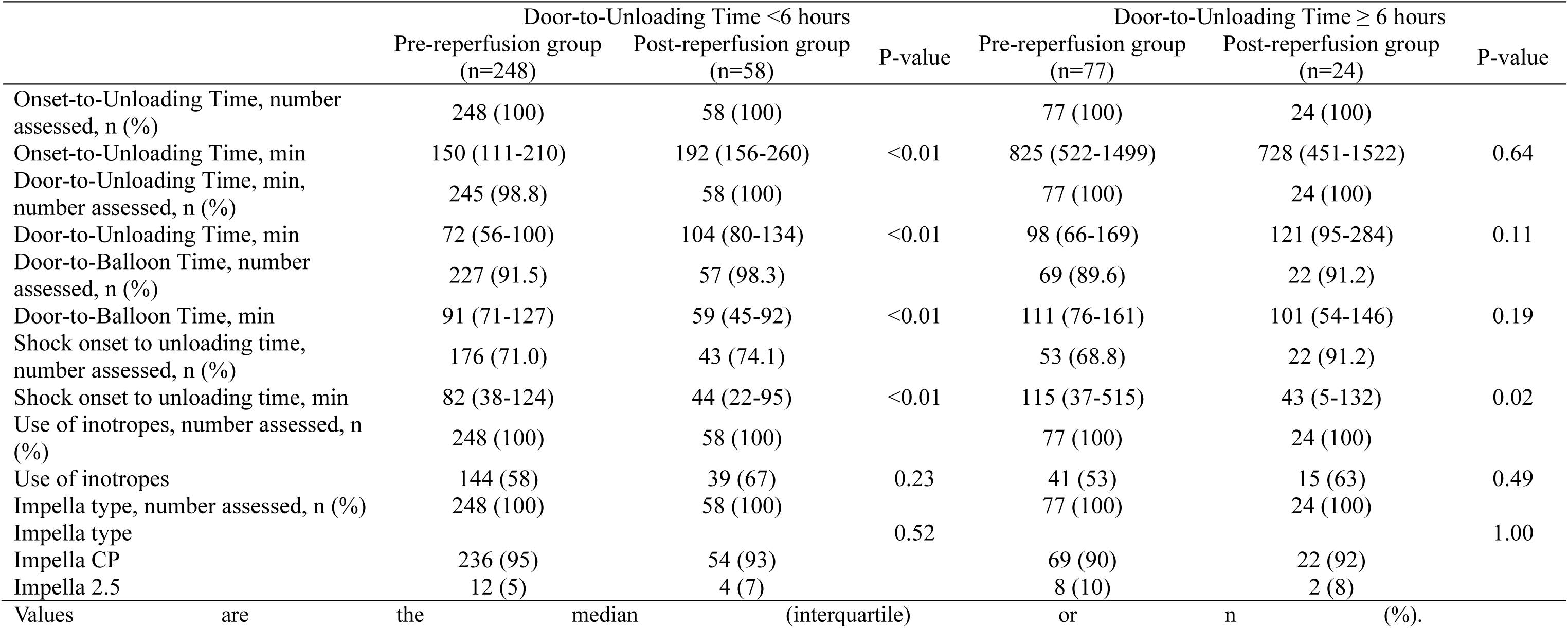
Procedural characteristics.

|  | Door-to-Unloading Time <6 hours |  |  | Door-to-Unloading Time ≥ 6 hours |  |  |
| --- | --- | --- | --- | --- | --- | --- |
|  | Pre-reperfusion group<br>(n=248) | Post-reperfusion group<br>(n=58) | P-value | Pre-reperfusion group<br>(n=77) | Post-reperfusion group<br>(n=24) | P-value |
| Onset-to-Unloading Time, number assessed, n (%) | 248 (100) | 58 (100) |  | 77 (100) | 24 (100) |  |
| Onset-to-Unloading Time, min | 150 (111-210) | 192 (156-260) | <0.01 | 825 (522-1499) | 728 (451-1522) | 0.64 |
| Door-to-Unloading Time, min, number assessed, n (%) | 245 (98.8) | 58 (100) |  | 77 (100) | 24 (100) |  |
| Door-to-Unloading Time, min | 72 (56-100) | 104 (80-134) | <0.01 | 98 (66-169) | 121 (95-284) | 0.11 |
| Door-to-Balloon Time, number assessed, n (%) | 227 (91.5) | 57 (98.3) |  | 69 (89.6) | 22 (91.2) |  |
| Door-to-Balloon Time, min | 91 (71-127) | 59 (45-92) | <0.01 | 111 (76-161) | 101 (54-146) | 0.19 |
| Shock onset to unloading time, number assessed, n (%) | 176 (71.0) | 43 (74.1) |  | 53 (68.8) | 22 (91.2) |  |
| Shock onset to unloading time, min | 82 (38-124) | 44 (22-95) | <0.01 | 115 (37-515) | 43 (5-132) | 0.02 |
| Use of inotropes, number assessed, n (%) | 248 (100) | 58 (100) |  | 77 (100) | 24 (100) |  |
| Use of inotropes | 144 (58) | 39 (67) | 0.23 | 41 (53) | 15 (63) | 0.49 |
| Impella type, number assessed, n (%) | 248 (100) | 58 (100) |  | 77 (100) | 24 (100) |  |
| Impella type |  |  | 0.52 |  |  | 1.00 |
| Impella CP | 236 (95) | 54 (93) |  | 69 (90) | 22 (92) |  |
| Impella 2.5 | 12 (5) | 4 (7) |  | 8 (10) | 2 (8) |  |

1 Values are the median (interquartile) or n (%).

### Survival rate and factors for survival in the onset-to-unloading time <6 hours cohort

A Kaplan-Meyer curve demonstrated the 80-day survival rate in the onset-to-unloading time <6 hours cohort (Figure 2A). Patients with pre- and post-reperfusion unloading had similar 80-day survival rates (85.3% vs. 91.2%, p=0.38). A univariable analysis revealed that the 80-day survival was related to the age, systolic blood pressure before the Impella insertion, laboratory data including the lactate dehydrogenase, creatinine, aspartate aminotransferase, alanine aminotransferase, creatine kinase, and C-reactive protein, door-to-unloading time, door-to-balloon time, and dyslipidemia (Table 3). Pre-reperfusion use of Impella was not associated with survival (hazard ratio 1.516 [95% confidence interval 0.591-3.892], p=0.39). After a multivariable analysis, the age (hazard ratio 1.054 [95% confidence interval 1.011-1.100], p=0.01), systolic blood pressure (hazard ratio 0.985 [95% confidence interval 0.971-0.999], p=0.04) and creatinine (hazard ratio 1.497 [95% confidence interval 1.231-1.819], p<0.01) were detected as independent predictors of survival.

**Figure 2.**
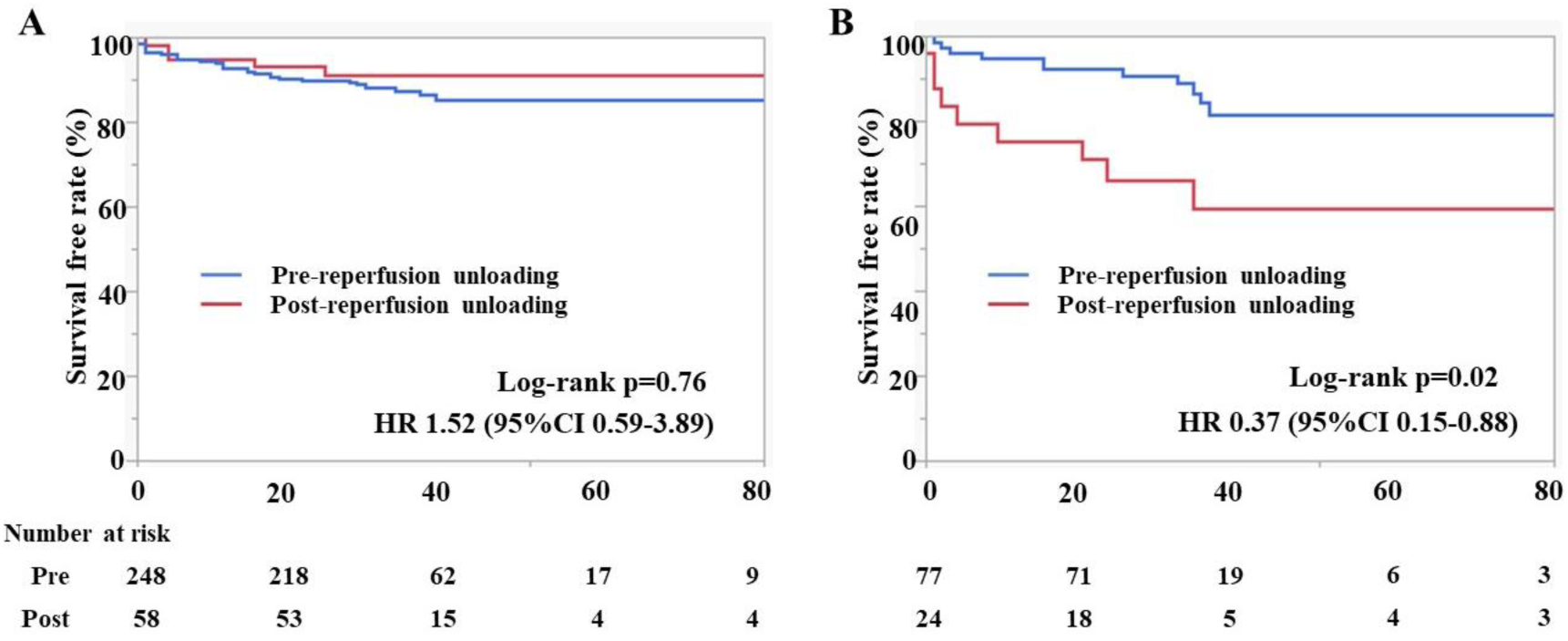
The 80-day survival rates and independent predictors of survival according to the onset-to-unloading time Kaplan-Meyer curves demonstrating the 80-day survival rates in the onset-to-unloading time <6 hours cohort (Figure A) and in the onset-to-unloading time ≥6 hours cohort (Figure B).

**Table 3.** Predictors of survival in the cohort with an onset-to-unloading time <6 hours.

|  | Univariable |  | Multivariable |  |
| --- | --- | --- | --- | --- |
|  | Hazard ratio | P value | Hazard ratio | P value |
| Age, year | 1.050 (1.018-1.084) | <0.01 | 1.054 (1.011-1.100) | 0.01 |
| Male, n (%) | 0.535 (0.253-1.135) | 0.10 |  |  |
| Body mass index, kg/m<sup>2</sup> | 1.028 (0.935-1.128) | 0.57 |  |  |
| Heart rate before Impella Insertion, bpm | 1.000 (0.987-1.012) | 0.98 |  |  |
| Systolic blood pressure before Impella insertion, mmHg | 0.989 (0.977-0.999) | 0.049 | 0.985 (0.971-0.999) | 0.04 |
| Diastolic blood pressure before Impella insertion, mmHg | 0.985 (0.968-1.0002) | 0.07 |  |  |
| Ejection fraction, % | 0.961 (0.921-1.0003) | 0.06 |  |  |
| Lactate, mmol/l | 1.006 (0.965-1.022) | 0.59 |  |  |
| Lactate dehydrogenase, U/L | 1.001 (1.001-1.001) | <0.01 | 1.002 (0.99991-1.004) | 0.06 |
| Creatinine, mg/dl | 1.264 (1.062-1.435) | <0.01 | 1.497 (1.231-1.819) | <0.01 |
| Total bilirubin, mg/dl | 1.600 (0.694-2.984) | 0.20 |  |  |
| Albumin, g/dl | 0.534 (0.290-1.023) | 0.05 |  |  |
| Aspartate aminotransferase, U/L | 1.002 (1.001-1.004) | <0.01 | 0.997 (0.983-1.011) | 0.68 |
| Alanine aminotransferase, U/L | 1.002 (1.000-1.004) | 0.03 | 1.001 (0.983-1.019) | 0.94 |
| Creatine phosphokinase, U/L | 1.001 (1.000-1.001) | <0.01 | 1.001 (0.998-1.002) | 0.34 |
| C-reactive protein, mg/dl | 1.098 (0.980-1.184) | 0.045 | 0.934 (0.752-1.161) | 0.54 |
| Onset-to-Unloading Time, min | 1.212 (0.942-1.545) | 0.13 |  |  |
| Door-to-Unloading Time, min | 1.032 (1.006-1.049) | <0.01 | 0.9997 (0.996-1.003) | 0.86 |
| Door-to-Balloon Time, min | 1.030 (1.002-1.048) | <0.01 | 1.001 (0.998-1.004) | 0.59 |
| Shock onset to unloading time, min | 0.962 (0.713-1.007) | 0.74 |  |  |
| Hypertension | 0.769 (0.665-2.542) | 0.44 |  |  |
| Dyslipidemia | 0.384 (0.192-0.768) | <0.01 | 0.600 (0.271-1.335) | 0.21 |
| Diabetes mellitus | 1.187 (0.615-2.292) | 0.61 |  |  |
| Chronic kidney disease | 1.833 (0.925-3.629) | 0.08 |  |  |
| Pre-reperfusion use of impella | 1.516 (0.591-3.892) | 0.39 |  |  |
| Impella CP | 0.967 (0.233-4.029) | 0.96 |  |  |
| Out-hospital arrest | 0.878 (0.270-2.859) | 0.83 |  |  |
| Use of inotropes | 1.427 (0.717-2.841) | 0.31 |  |  |

2 Values are the hazard ratio (95% confidence interval).

### Survival rate and factors for survival in the onset-to-unloading time ≥6 hours cohort

Patients with pre-reperfusion unloading had a significantly higher 80-day survival rate than the patients with post-reperfusion unloading (81.6% vs. 59.5%, p=0.02) (Figure 2B). The hazard ratio of survival in patients who underwent pre-reperfusion unloading versus post-reperfusion unloading was 0.37 (95% confidence interval 0.15-0.88). While a univariable analysis demonstrated that the age, gender, diastolic blood pressure before the Impella insertion, lactate, albumin, and pre-reperfusion use of Impella were associated with survival (Table 4), a multivariable analysis revealed that the independent predictors of survival were the age (hazard ratio 1.137 [95% confidence interval 1.030-1.295], p=0.02), diastolic blood pressure (hazard ratio 0.959 [95% confidence interval 0.927-0.995], p=0.02), and pre-reperfusion use of Impella (hazard ratio 0.184 [95% confidence interval 0.045-0.746], p=0.02).

**Table 4.** Predictors of survival in the cohort with an onset-to-unloading time ≥6 hours.

|  | Univariable |  | Multivariable |  |
| --- | --- | --- | --- | --- |
|  | Hazard ratio | P value | Hazard ratio | P value |
| Age, year | 1.150 (1.075-1.248) | <0.01 | 1.137 (1.030-1.295) | 0.02 |
| Male, n (%) | 0.353 (0.140-0.893) | 0.03 | 3.470 (0.436-27.587) | 0.24 |
| Body mass index, kg/m<sup>2</sup> | 0.968 (0.848-1.085) | 0.61 |  |  |
| Heart rate before Impella Insertion, bpm | 0.997 (0.976-1.019) | 0.85 |  |  |
| Systolic blood pressure before Impella insertion, mmHg | 0.989 (0.971-1.006) | 0.22 |  |  |
| Diastolic blood pressure before Impella insertion, mmHg | 0.978 (0.960-0.998) | 0.02 | 0.959 (0.927-0.995) | 0.02 |
| Ejection fraction, % | 0.981 (0.924-1.031) | 0.48 |  |  |
| Lactate, mmol/l | 1.192 (1.020-1.379) | 0.02 | 1.063 (0.858-1.313) | 0.56 |
| Lactate dehydrogenase, U/L | 1.000 (0.999-1.001) | 0.91 |  |  |
| Creatinine, mg/dl | 0.969 (0.633-1.237) | 0.84 |  |  |
| Total bilirubin, mg/dl | 1.325 (0.657-2.028) | 0.30 |  |  |
| Albumin, g/dl | 0.436 (0.198-0.961) | 0.04 | 0.520 (0.148-2.030) | 0.32 |
| Aspartate aminotransferase, U/L | 1.000 (0.997-1.001) | 0.95 |  |  |
| Alanine aminotransferase, U/L | 1.000 (0.997-1.001) | 0.85 |  |  |
| Creatine phosphokinase, U/L | 1.000 (1.000-1.0004) | 0.52 |  |  |
| C-reactive protein, mg/dl | 0.995 (0.878-1.094) | 0.93 |  |  |
| Onset-to-Unloading Time, min | 0.998 (0.988-1.003) | 0.57 |  |  |
| Door-to-Unloading Time, min | 1.000 (0.983-1.0009) | 0.99 |  |  |
| Door-to-Balloon Time, min | 1.002 (0.987-1.011) | 0.67 |  |  |
| Shock onset to unloading time, min | 0.993 (0.932-1.004) | 0.69 |  |  |
| Hypertension | 0.529 (0.219-1.278) | 0.16 |  |  |
| Dyslipidemia | 0.660 (0.273-1.600) | 0.36 |  |  |
| Diabetes mellitus | 1.714 (0.719-4.084) | 0.22 |  |  |
| Chronic kidney disease | 2.168 (0.913-5.148) | 0.08 |  |  |
| Pre-reperfusion use of impella | 0.368 (0.154-0.882) | 0.03 | 0.184 (0.045-0.746) | 0.02 |
| Impella CP | 0.969 (0.233-4.029) | 0.96 |  |  |
| Out-hospital arrest | 2.404 (0.704-8.210) | 0.16 |  |  |
| Use of inotropes | 1.427 (0.717-2.841) | 0.31 |  |  |

2 Values are the hazard ratio (95% confidence interval).

### Major adverse events

The clinical event data were available for all patients. Hemorrhage or hematoma was more frequently observed in patients with pre-reperfusion Impella than in those with post-reperfusion Impella in the cohort with a door-to-unloading time <6 hours (p=0.04). The other events were similarly seen in each group in both cohorts (Table 5).

**Table 5.**
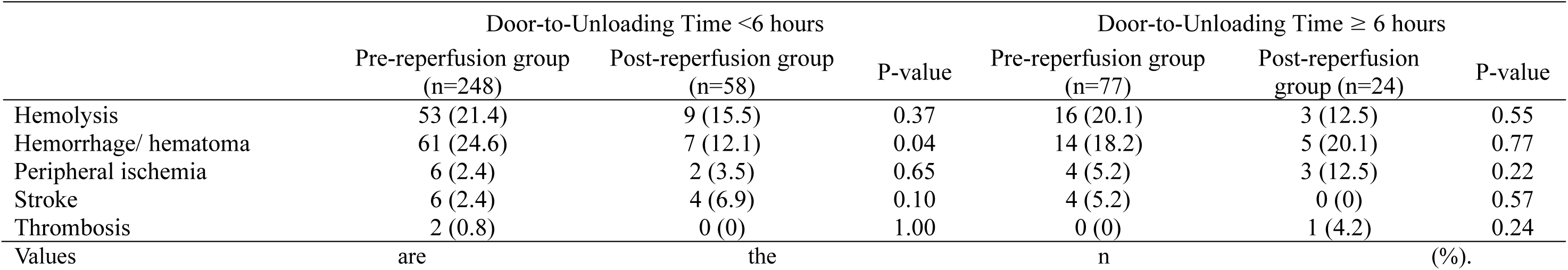
Major adverse events.

## Discussion

The present study is the first clinical large-scale report to demonstrate the differences in the survival outcomes associated with LV unloading using Impella prior to PCI in patients with STEMI, based on the onset-to-unloading time. The main findings of the study included the following: (1) pre-reperfusion LV unloading with Impella significantly reduced the mortality rate compared with post-reperfusion unloading in the onset-to-unloading ≥6 hours cohort, while pre- and post-reperfusion unloading in patients with STEMI resulted in the same 80-day survival rate, and the timing of the Impella insertion was not associated with the survival in the onset-to-unloading <6 hours cohort; (2) pre-reperfusion use of Impella, high age, and low diastolic blood pressure were independent factors contributing to the survival in the onset-to-unloading ≥6 hours cohort. High age, low systolic blood pressure, and high creatinine level were independent predictors of survival in the onset-to-unloading <6 hours cohort.

STEMI is generally caused by a sudden occlusion of an epicardial artery and the myocardium distal to the occlusion site becomes ischemic. The unrelieved ischemic myocardium suffers permanent damage and the hypoperfused myocardial zone is known as the area at risk (AAR). If the occluded artery is not reperfused, most of the AAR results in necrosis ^12, 13^. The ischemia injury arises from the subendocardium and progresses into the subepicardium when the occlusion time is longer than 20 minutes. The irreversible ischemia injury increases in size gradually over time ^14^. Approximately 30-50% of the AAR remains viable after 4 to 6 hours from the onset based on the amount of the salvaged myocardium at the time of reperfusion ^12, 15, 16^. Even after 12 hours from the onset, reperfusion treatment can significantly reduce the infarct size ^17^. The development of the reperfusion therapy including a progressive reduction in the time between the STEMI diagnosis and reperfusion dramatically improved the survival rate over several decades ^18^. Therefore, primary PCI as early as possible is recommended as a standard therapy in the current guidelines to reduce the ischemia injury ^4, 5^. While primary PCI is the most effective strategy for reducing the infarct size, reperfusion injury can paradoxically reduce the benefit of the reperfusion therapy. The reperfusion injury is defined as myocardial injury caused by the restoration of blood flow after ischemia and leads to the death of the cardiomyocytes that are viable immediately before reperfusion ^19^. The final infarct size is determined by both the ischemia and reperfusion injury (Figure 3A) ^13^. The infarct size has been shown to be strongly associated with all-cause mortality and hospitalization for heart failure even in the contemporary primary PCI era ^20^. Therefore, the infarct size is one of the main factors affecting the clinical outcomes after STEMI.

**Figure 3.**
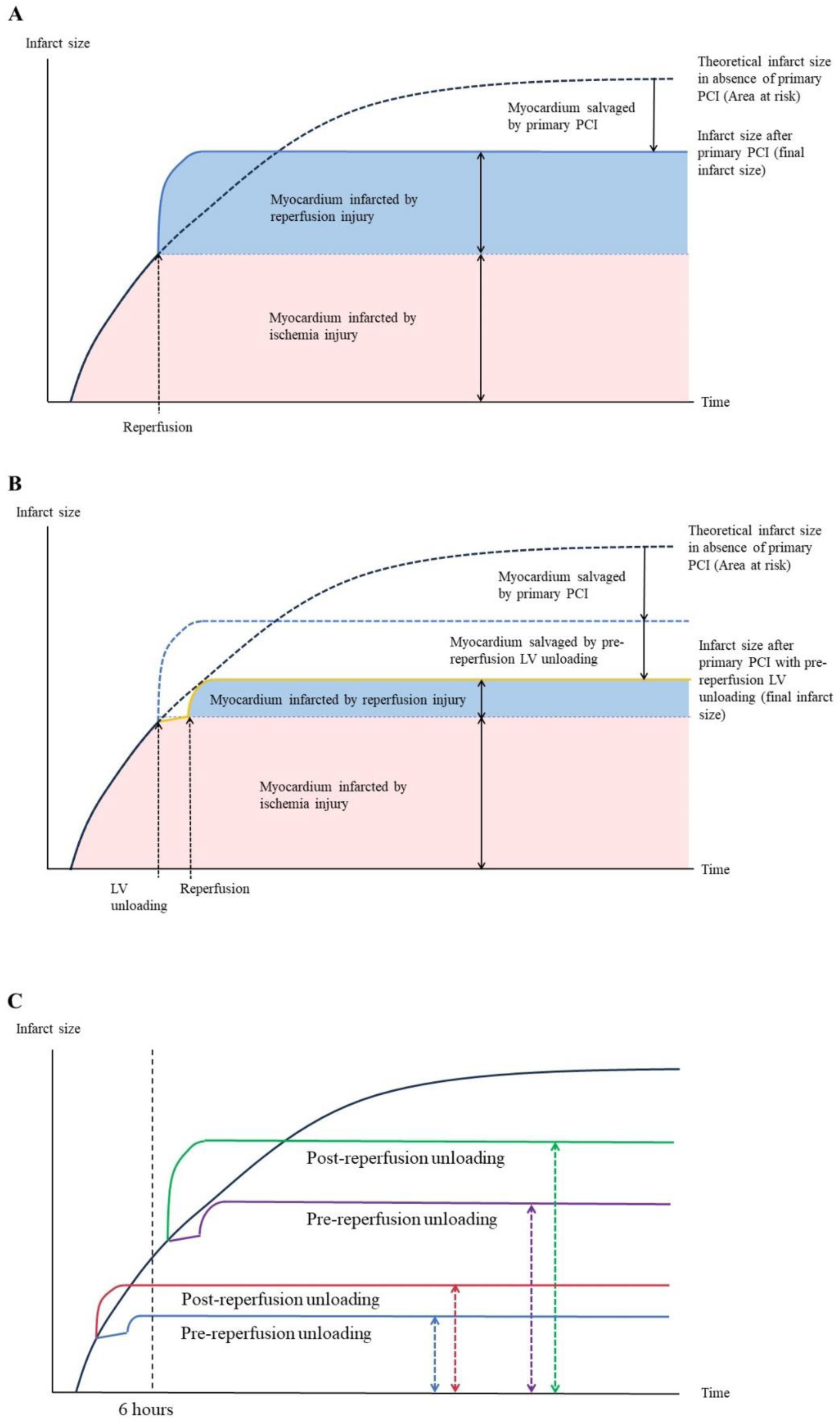
Theoretical infarction size Figure A presents the infarct size after primary percutaneous coronary intervention (PCI) over time after the onset of symptoms in patients with ST-segment-elevation myocardial infarction (STEMI). The red area indicates infarcted myocardium due to ischemia injury and the blue area depicts myocardium infarcted by reperfusion injury. The black dashed line demonstrates the theoretical infarct size in the absence of primary PCI. Figure B illustrates the final infarct size after primary PCI with left ventricle (LV) unloading prior to reperfusion using Impella. The blue dotted line describes the infarct size when treated with only primary PCI. The yellow line depicts the change in the infarct size over time after LV unloading and reperfusion. LV unloading can increase the collateral flow and reduce the LV oxygen consumption, leading to a decrease in the progression of ischemia injury. Furthermore, activating a cardioprotective program reduces the reperfusion injury. Finally, the infarct size decreased with pre-reperfusion LV unloading. Figure C demonstrates the theoretical infarct size in the pre- and post-reperfusion unloading groups in each cohort. The blue and red lines indicate the infarct size of STEMI treated with primary PCI and pre- or post-reperfusion unloading in the cohort with an onset-to-unloading time <6 hours. The purple and green lines depict the theoretical infarct size that resulted after the primary PCI and pre- or post-reperfusion unloading in the cohort with an onset-to-unloading time ≥6 hours.

The Impella has not only an effect of cardiac support for cardiogenic shock but a potential to reduce the infarct size by the following effects; 1) an increase in the collateral flow, which can reduce the ischemia injury through a decrease in the ARR ^21^; 2) activating a cardioprotective program and preserving the mitochondrial structure, which can decrease the reperfusion injury ^6, 7^; and 3) a reduction in the LV oxygen consumption, which may reduce both the ischemia and reperfusion injury ^21, 22^. Several animal studies have shown that the effects of a decrease in the infarction size are maximally obtained when LV unloading using Impella is started before reperfusion despite a delayed onset-to-reperfusion time ^6, 22^ (Figure 3B).

The survival rate in the pre-reperfusion unloading group was significantly higher than that in the post-reperfusion unloading group beyond 6 hours after the onset, while a survival rate of 80 days was similar within 6 hours after the onset of the symptoms. This may indicate that the time from the onset should be a critical factor of the infarction size and it is possible that the infarct size in both groups was not large enough to significantly affect the short-term survival rate in the early phase after the onset. On the other hand, the infarct size progressively increases over time and the reduced infarct size by pre-reperfusion LV unloading may have a significant impact on the short-term survival rate in the late phase (Figure 3C). However, the long-term clinical outcomes including death and hospitalization for heart failure may differ between the two groups even in the early phase after the onset, and further investigation is warranted. In addition, the results confirmed that the delayed reperfusion due to pre-reperfusion LV unloading did not result in adverse clinical effects, even during the early phase, despite a significantly longer door-to-balloon time in the pre-reperfusion unloading group as compared with the post-reperfusion unloading group. The result indicated that we could perform pre-reperfusion unloading safely even in the early phase after the onset.

Impella use prior to PCI was associated with an improved survival in several registries despite the lack of time from the onset ^8, 9^. The Uspella registry demonstrated that pre-PCI Impella use improved the discharge survival rate than post-PCI Impella use, with an odds ratio of 0.37 (95% CI: 0.19-0.72). The registry showed that 52.8% of the patients had a shock duration of more than 6 hours at baseline, indicating that the onset-to-unloading time should be greater than 6 hours in a larger study population ^8^. These characteristics of the study population could potentially lead to better survival outcomes with pre-reperfusion Impella use as compared with post-reperfusion Impella use. The “SHould we emergently revascularize Occluded Coronaries for cardiogenic shock” (SHOCK) trial reported that the systolic and diastolic blood pressures were associated with in-hospital mortality in patients with cardiogenic shock accompanied by acute myocardial infarction ^23^. The National Cardiogenic Shock Initiative study demonstrated that age ≥ 70 years and creatinine ≥ 2mg/dl were predictors of in-hospital mortality ^24^. This study revealed that high age and low blood pressure were predictors of mortality in both the early and late phases after the onset. Furthermore, a high creatinine value was identified to be an independent predictor of mortality in the early phase. Those results were consistent with the results of the previous studies. While the age, blood pressure, and creatinine value are patient conditions and difficult to manage, the timing of starting the LV unloading mainly depends on the operators’ discretion and is comparatively easy to control. Pre-reperfusion LV unloading should be considered to improve the short-term survival especially when more than 6 hours have passed since the onset of symptoms.

## Limitations

This study was conducted using an observational registry. Since patients with no data regarding the onset-to-unloading time, Impella support timing, and survival were excluded, there might have been bias in the study population. Additionally, several patient characteristics and laboratory data were lacking.

## Conclusions

Pre-reperfusion LV unloading can be a crucial treatment to improve the short-term survival rate when the onset-to-LV unloading is more than 6 hours.

## Data Availability

Raw data were generated at the Japan VAD Council, IMPELLA Committee. Derived data supporting the findings of this study are available from the corresponding author on request.

## Acknowledgements

The authors thank Mr. John Martin for his linguistic assistance with this manuscript.

## Sources of Funding

This study was funded by a donation from Abiomed Japan and self-financing by the Japan VAD Council, IMPELLA Committee.

## Disclosures

Masami Nishino receives donations from Abbott Medical Japan, Boston Scientific Japan, Medtronic, Japan Lifeline. However, the donation is not for this study. The other authors report no conflict.

## Clinical Perspectives

### What is new?

Patients with pre-reperfusion unloading had a significantly higher 80-day survival rate than the patients with post-reperfusion unloading, and pre-reperfusion use of Impella was an independent factor of survival in the onset-to-unloading time ≥6 hours cohort.
The timing of starting LV unloading did not affect the short-term survival rate in the cohort with an onset-to-unloading time <6 hours.

### What are the clinical implications?

It is clinically important to verify which patient groups will benefit from the pre-reperfusion LV unloading with Impella. Pre-reperfusion LV unloading could be a crucial treatment to improve the short-term survival rate when the onset-to-LV unloading was more than 6 hours.

## Non-standard Abbreviations and Acronyms

AAR: area at risk
LV: left ventricle
PCI: percutaneous coronary intervention
STEMI: ST-segment-elevation myocardial infarction

## Notes

### Clinical Trial

The Universal Hospital Medical Information Network (UMIN) Clinical Trial Registry (ID: UMIN 000033603)

### Author Declarations

The study was approved by the Central Institution Review Board at Osaka University.

